# Mathematical Relationship between Reproduction Number and Epidemic Curve of Daily Cases

**DOI:** 10.1101/2021.01.24.21250405

**Authors:** Bruno Caudana

## Abstract

The strict mathematical relationship between *R*_*t*_ and the curve of daily cases *f* (*t*) is shown. Up to date and statistically robust *R*_*t*_ from the curve of daily cases can be estimated as soon as new cases are added to the curve. That is equivalent to estimating *R*_*t*_ by averaging all detected cases of infection, without any distortion induced by the difficulty of following and weighting trees of secondary cases from original ones, and without needing to wait for secondary cases to manifest infection. With this method, if *R*_*t*_ scaled numbers are of interest, only the average duration of subjects’ infectivity has to be estimated directly, but independently of linking secondary cases to primary ones. A new index, *Instantaneous Reproduction Number R*_*ist*_ is introduced, which is mathematically tied to *R*_*t*_ and does not depend on the duration of infectivity of subjects. *R*_*ist*_, *R*_*t*_ and the doubling/halving time of the epidemic may be estimated by simple computations at the very detection time of new daily cases. Any smoothed curve of daily cases gives smooth *R*_*t*_ and *R*_*ist*_. No phase lag on *R*_*t*_ estimate is introduced by this method.

## Motivation for this method

During the first phase of COVID19 outbreak, while tinkering with a diffusion-saturation model to fit epidemic data [7], I encountered estimations of *R*_*t*_ and *R*_0_ which where incompatible with the observed doubling time of daily cases and the location in time of peaks. So, I began to think on the subject.

It seems that *R*_*t*_ was defined from the epidemiological point of view with the assumption in mind that an epidemic can be characterized by a somewhat stable relationship between a pathogen and its infectable host, with the intent of predicting the evolution of an outbreak. Which does not seem to work properly.

In fact, the initial susceptibility of a population of hosts is always unknown, because unknown is the reaction of the immune system spectrum and history of a population to a new pathogen or to similar ones as biochemical targets.

Besides that, both pathogen and host can have this relationship modified via several options (the decreasing susceptibility of the host population due to the spreading of the epidemic that saturates the susceptibility of a population or sub-population; the reaction of immune systems; the reactive behaviours of host and pathogen populations; the mobility pattern of vector particles, like infecting particles in turbulent air; etc).

This writing shows how *R*_*t*_ definition is strictly tied to the curve of daily cases by mathematical equations. The two are essentially the same thing expressed with different words. *R*_*t*_ is a sort of first derivative of the curve of daily cases with respect to time *t*.

The difficulty of directly estimating *R*_*t*_ in a reliable way is the same as predicting the evolution of an epidemic in a reliable way. Indeed even much harder, since one has to face the further uncertainty of estimating and weighting trees of secondary cases implied by this process. It is very similar to estimating the space traveled by measuring acceleration with very inaccurate accelerometers, but very much harder and error prone.

The excellent articles by Cori, et al. [1] and Dietz [2] clearly show this difficulty.

### Epidemiological definition of R_*t*_

The epidemiological definition of *R*_*t*_ states:

*R*_*t*_ *is the number of secondary infections caused by a single case of disease during its period of infectivity in a completely susceptible population, on average*.

(see: [3], [1], [2] and many other sources.)

According to this epidemiological definition, *R*_*t*_ is analogous to the multiplier of the initial unit capital after 1 period, in a compound capitalization process.

This analogy allows the estimation of *R*_*t*_ from the epidemic curve of daily cases by introducing the concept of *Instantaneous Reproduction Number R*_*ist*_, similar to the instantaneous capitalization rate in actuarial mathematics.

### Definition of R_*ist*_

The epidemiological definition of *R*_*t*_ indicates an exponential expansion. So does *R*_0_, as the limit to the beginning of an epidemic of an uninfected population. An infected individual, after his period of infectious capacity, will have infected a new infected plus (or minus) a number of new infected, on average. This is equivalent to the amount of a compound capitalization at the interest rate *r*, where *R*_*t*_ is the amount after period 1. In general:

- after period 1: 1 · (1 + *r*) = *R*_*t*_;
- after period 2: 1 · (1 + *r*) · (1 + *r*) = 1 · (1 + *r*)^2^;
- after period *p*: 1 ·(1 + *r*)^*p*^.

To obtain which interest rate *r* should be used for a continuous compound capitalization of *n* fractions of a period that gives the amount *R*_*t*_ after 1 period, we can write as follows:

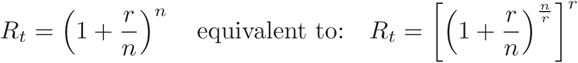

Passing to the limit for *n* → ∞, and noting that 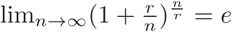, we get:

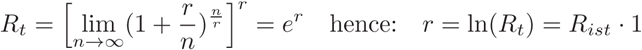

In other terms, *r* is the exponent to be given to *e* to obtain *R*_*t*_ after a period of infectious duration equal to 1.

If we want to express *R*_*ist*_ in a unit of time *g*_*i*_ other than the dimensionless unit period, for example the days (or hours) with which we measure the duration of the infectivity period of an infectious subject and with which we measure the progress of the epidemic, we can write:

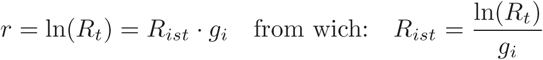

In this way we have the parameter *R*_*ist*_ which characterizes the exponential growth (as per the definition of *R*_*t*_) at the point in time *t* that the increase (or decrease) of daily cases generates.

### Connecting R_*ist*_ to the epidemic curve of daily cases

Whenever an exponential function *y* = *e*^*ax*^ is represented in logarithmic scale ln(*y*) = *ax*, it becomes a straight line. Its shape factor *a* becomes the slope of the straight line (the angular coefficient).

If we represent the curve of the daily cases *f* (*t*) in logarithmic scale *h*(*t*) = ln(*f* (*t*)), the slope of the tangent of *h*(*t*) at point *t* is the slope *R*_*ist*_, corresponding to the exponential growth of the epidemiological definition of the effective reproduction number *R*_*t*_, represented in logarithmic scale, at time *t*, and scaled in time units of the curve of daily cases. But the tangent of *h*(*t*) at point *t* is also the first derivative of *h*(*t*). That is:

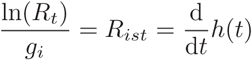

A different reasoning perhaps better illustrates the concept of estimating *R*_*t*_ from epidemic curves.

Since all infected have been infected by someone, *R*_*t*_ is basically the ratio between the daily cases at time *t* + 1 over the cases at time *t*, where 1 is the infecting period. Given the point *a* on the curve of daily cases that precedes the point *b*, then *R*_*t*_ = *b/a*.

Differentiating the curve of daily cases, expressed in logarithmic scale with base *e*, means making the difference between two values, spaced by a unitary period of time tending to zero, that is: ln(*b*) − ln(*a*). This expression is equivalent to doing ln(*b/a*), as those who have used slide rules [4] easily remember: ln(*b*) − ln(*a*) = ln(*b/a*).

By doing the inverse operation of extracting a logarithm from a number, i.e. raising the base of the logarithm to a power of the value of the logarithm in question, one obtains the ratio *b/a* in the scale of daily cases of infection: *e*^ln(*b/a*)^ = *b/a*.

This ratio represents the rate of increase (if *>* 1), or decrease (if *<* 1), of the infections averaged over all the infections observed, including all the information on the overall average resistance to the spread of the infection that may have formed meanwhile, for any known or unknown reason it was formed. It also takes in properly weighted account all the overlaps of the infection trees defined by *R*_*t*_, and of the hosts’ varying susceptibility.

Furthermore, the value obtained in this way is a very accurate value of *R*_*t*_ acting at current time of *b*, that is, at the very moment in which the current value of the infected cases is known. The passage to the limit of a period that tends to the instant, implicit in the differentiation operation with respect to *t*, allows to have a curve of *R*_*t*_ trend that is always updated in real-time.

According to the epidemiological definition of *R*_*t*_, we have the following correspondence of classical outstanding cases, direct consequence of the epidemiological definition:

*R*_*ist*_ *>* 0 : when the daily cases increase and the epidemic is expanding; therefore the associated 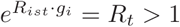.

*R*_*ist*_ = 0 : when the daily cases remain constant and the epidemic is stationary therefore the associated 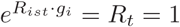. In this case the curve of daily cases has a minimum or a maximum; *R*_*ist*_ crosses 0; *R*_*t*_ crosses 1.

*R*_*ist*_ *<* 0 : when the daily cases decrease and the epidemic is contracting therefore the associated 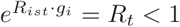.

Since these outstanding cases derive from the epidemiological definition of *R*_*t*_, they also are criterion for evaluating the correct estimate of *R*_*t*_. A contrasting value of *R*_*t*_ respect to the epidemic curve is also an indication that *R*_*t*_ or the epidemic curve are wrong.

### Summary of conversion formulas

The curve of daily cases *f* (*t*) expressed in logarithmic scale with base *e* is obviously given by:

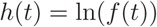

*R*_*ist*_ is given by the first derivative (numerically or analytically determined) of any smoothed curve of daily cases, given in logarithmic scale with base *e*:

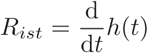

Please notice that if we have a smoothing procedure of the curve of daily cases that introduces any phase lag, as we have using mobile averages or FIR/IIR filters, we will have the same phase lag in the estimation of *R*_*ist*_ and *R*_*t*_. Otherwise if we have some form of static averaging, as using some least squares fitting procedure, no phase lag is introduced.

*R*_*t*_ is given by:

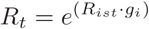

*R*_*ist*_ is also equivalen to:

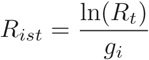

The doubling or halving time of infection *g*_*d*?*h*_ is given by imposing 2.0 as *R*_*t*_ and computing the number of resulting days (negative numbers represent halving time):

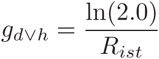

### Some generable charts

The following charts show how *R*_*t*_ may be calculated from a fitting of the curve of cumulative cases. Obviously, the curve of daily cases is the first derivative of cumulative cases. The second derivative is the one obtained by differentiating the curve of daily cases taken in logarithmic scale to generate *R*_*ist*_. Taken together, they form *a sort of* derivative of order 2 of the curve of cumulative cases.

The fitting is primarily done on cumulative cases because they automatically compensate some kind of errors (for example: a missed case one day may be detected in the following days, etc.). Model and fitting techniques [7] used for the following figures are outside the scope of this writing. Here the model is simply used as source of a smoothed daily data set. The other formulas used to generate the following charts are summarized in the section above. The data source used for this fitting is the COVID-19 official one for Italy [6].

**Figure 1.**
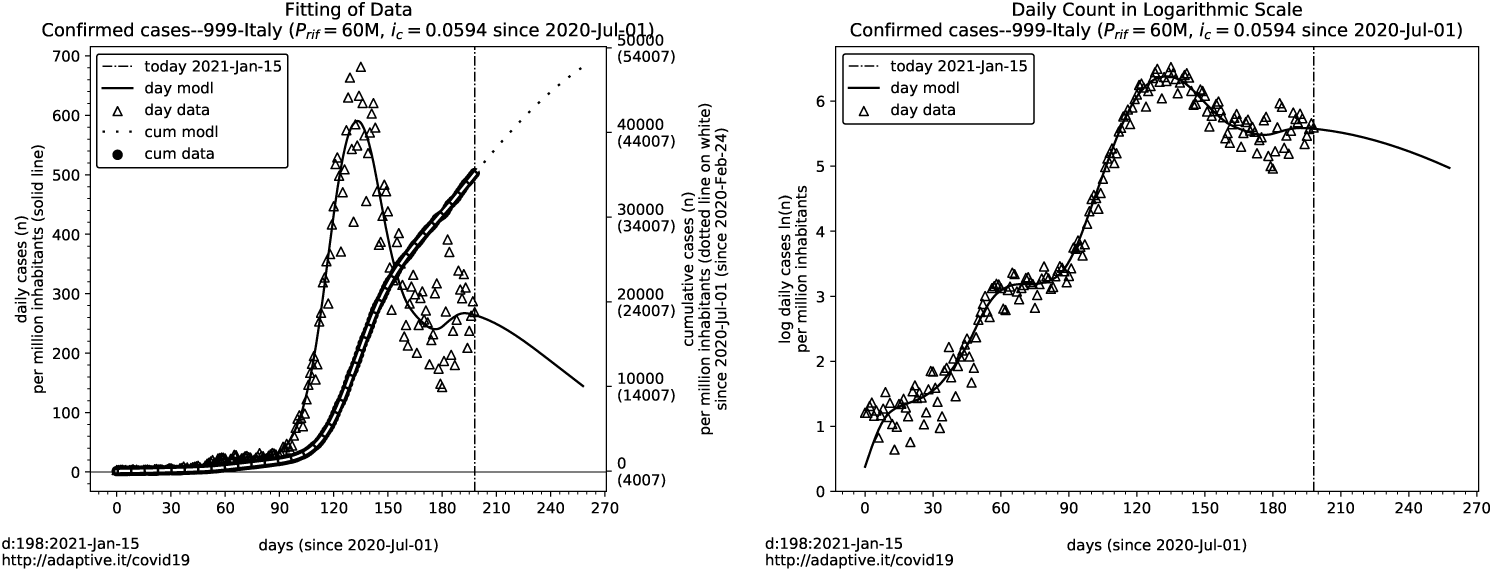
a) Data fit with an adaptive diffusion-saturation model on cumulative cases.b)Daily cases smoothed by fitting, in logarithmic scale with base *e* used for generating *R*_*t*_ and *R*_*ist*_. (Model and fitting techniques used for these charts are outside the scope of this writing. Data source: [6]. Fitting model and details: [7])

**Figure 2.**
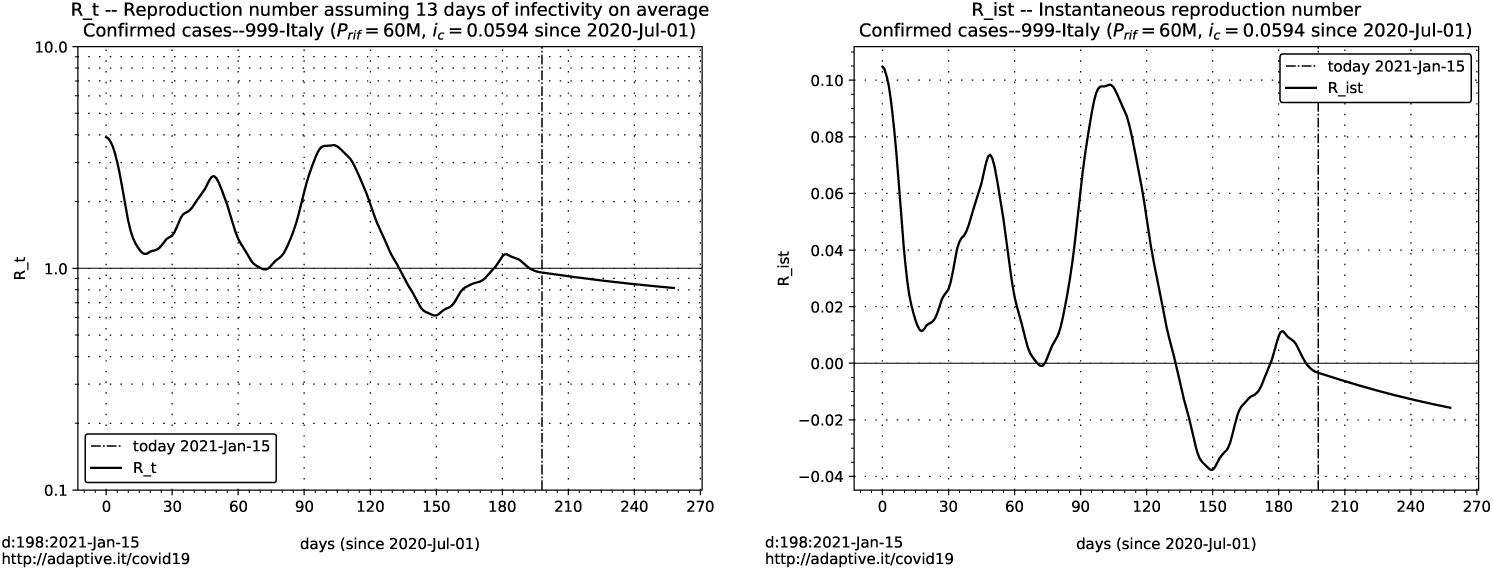
*R*_*t*_ and *R*_*ist*_ computed from the fitted curve of daily cases in logarithmic scale

## Conclusion

*Reproduction number R*_*t*_ and *epidemic curve of daily cases* are essentially the same thing expressed with different words. The conversion formulas between the two are shown.

The strict mathematical relationship between the *Reproduction Number R*_*t*_ and the *epidemic curve of daily cases* allows to substitute complex and error prone statistical estimations with deterministic computations from the count of daily cases. It is still necessary to directly estimate the average duration of the infectivity of subjects, but independently of linking secondary cases to primary ones. Moreover, the obtained estimation of *R*_*t*_ is done averaging all detected cases, instead of a possibly deviated sample of them..

The method also completely eliminates, by mathematics, the uncertainty introduced by the process of identifying secondary cases from primary ones along overlapping trees of infectious relationships. Since all detected daily cases are infected by someone, the resulting curve of daily cases is the perfectly balanced action of *R*_*t*_, and it is detected in full. This method automatically incorporates that balance.

Also, the *R*_*t*_ estimate performed by this method is always up to date, as soon as new cases are added to the epidemic curve, without the need to wait for secondary cases to manifest after the period of infection.

Just a glance at the dispersion of an ample set of daily data around a good fitting of these data let easily imagine how difficult and unreliable could be any attempt to directly estimate a trend of the epidemic from small samples of cases and relying on considerations of the spread of these samples over overlapping trees of secondary cases. But this is exactly what the epidemiological definition of *R*_*t*_ asks to do.

A more general conclusion may be drawn by observing how different is the evolution of an epidemic of the same pathogens in different regions or times. The ample difference of *R*_*t*_ curves computed by this method on different zones [8] clearly shows that the dynamics of an epidemic seems to follow unpredictable and chaotic behavior, very much like weather.

We are used to think of populations involved in an epidemic as an isotropic material, like steel, which has equal behavior in all directions respect to stress and strain. SIR models and the epidemiological definition of *R*_*t*_ seem to implicitly assume this isotropy.

Perhaps, an epidemic may better be depicted as acting on many different relationship’s fabrics entangled together. A burst of infections occurs when two or more entangled fabrics mix and new connections merge in a new more extended fabrics–entangled fabrics which may be some in a stable infectious condition that eventually becomes saturated, and others not.

If this is a plausible landscape of a population’s infection, not every link in this entanglement of networks has the same infection capacity and not all nodes of these networks are isotropically connected.

In other words, there may be several networks that may have poor connections with each other, while having strong connection among the members of each network. For example, the network of families with children that go to the same school may have strong links between families of teachers and classmates, but may have weak connections with other unrelated networks of parents-children-teachers. Some of these networks may saturate eventually, while others may not have even been infected. The same thing happens with other types of relational networks. This is a very anisotropic environment.

This landscape shows a very challenging non linear object to investigate. Maybe, it has some emerging regularities at the macroscopic level, like sequences of overlapping sigmoidal shapes in the curve of cumulative cases, just as a complex sound emerges from the combination of elementary sinusoids. World [5] and regional [8] COVID-19 data of cumulative cases show everywhere overlapping sigmoids.

Maybe sophisticated tools, like the emerging field of “dynamical networks”, will provide interesting insights. However, to build reliable models based on the dynamics of networks is very difficult. The risk always is that the model needs a one-to-one mapping to the unknown phenomenon object of modelling, which leads to the conclusion that the proof may only be in the pudding.

## Data Availability

All the data used are publicly available.
The mathematical model used is made by the author.

https://github.com/pcm-dpc/COVID-19/

http://adaptive.it/covid19/

## Acknowledgments

Thanks to all those who taught me some mathematics and computing. Special thanks to prof. Roberto Merletti, prof. Giorgio Palù, prof. Fabio Truc, dr. Aldo Gotta, prof. Giulio Tarro and prof. Federico Butera who appreciated this work at various stages of development.

## References

1. Cori A, Ferguson NM, Fraser C, Cauchemez S. 2013 A New Framework and Software to Estimate Time-Varying Reproduction Numbers During Epidemics. American Journal of Epidemiology 178, 1505–1512. doi:10.1093/aje/kwt133

2. Dietz K. 1993 The estimation of the basic reproduction number for infectious diseases. Statistical Methods in Medical Research 2, 23–41. doi:10.1177/096228029300200103

3. Basic reproduction number. Wikipedia See https://en.wikipedia.org/wiki/Basic reproduction number

4. Slide rule. Wikipedia See https://en.wikipedia.org/wiki/Slide rule

5. Cumulative confirmed cases. Our World in Data See https://ourworldindata.org/coronavirus-data-explorer

6. 2020-2021 Protezione Civile Dati COVID-19 Italia. See https://github.com/pcm-dpc/COVID-19/

7. 2020-2021 Modello a diffusione-saturazione per andamento COVID-19. See http://adaptive.it/covid19

8. 2020-2021 Different curves in different zones at the same time with almost identical lockdowns and restraints. See http://www.adaptive.it/covid19/regions.php Sicily: http://adaptive.it/covid19/thiszoneit.php?zn=19Lombardy: http://adaptive.it/covid19/thiszoneit.php?zn=3SouthTyrol: http://adaptive.it/covid19/thiszoneit.php?zn=21

